# The Psychosis MRI Shared Data Resource (Psy-ShareD)

**DOI:** 10.1101/2024.10.03.24314828

**Authors:** Paul Allen, Mariana Zurita, Rubaida Easmin, Sara Bucci, Matthew J Kempton, Jack Rogers, Urvakhsh M Mehta, Philip K McGuire, Stephen M Lawrie, Heather Whalley, Ary Gadelha, Graham K Murray, Jane R Garrison, Sophia Frangou, Rachel Upthegrove, Simon L Evans, Veena Kumari, the Psy-ShareD Partnership

## Abstract

A key line of research in the field of schizophrenia and other psychotic disorders has been to investigate neuroanatomical markers, relative to healthy control groups. In recent decades, a large number of structural Magnetic Resonance Imaging (MRI) studies have been funded and undertaken, but their small sample sizes and heterogenous methods have led to inconsistencies across findings. To tackle this, efforts have been made to combine datasets across studies and sites. While notable recent multi-centre initiatives and both meta- and mega-analytical studies have progressed the field, efforts have generally been restricted to MRI scans in one or two illness stages which are not always representative of the diversity of patients who experience psychosis. Furthermore, access to these datasets is usually restricted to consortia members, often from high income countries. The Psychosis MRI Shared Data Resource (Psy-ShareD) is a new open access structural MRI data sharing partnership that will host pre-existing structural T1-weighted MRI data collected across multiple sites worldwide, including the Global South. MRI T1 data included in Psy-ShareD will be available in image and feature level formats, having been harmonised using state-of-the-art approaches. All T1 data will be linked to demographic and illness-related (diagnosis, symptoms, medication status) measures and in a number of datasets IQ and cognitive data, and medication history will also be available. Psy-ShareD will be free to access for all researchers. Comprehensive data catalogues, support and training resources will be available to facilitate use by early career researchers and build capacity in the field. We are actively seeking new collaborators to contribute further T1 data. Collaborators will benefit in terms of authorships, as all publications arising from Psy-ShareD will include data contributors as authors.

## 1 Introduction

Despite decades of research investment, schizophrenia spectrum disorders are still a leading cause of disability (Lopez et al., 2006; www.who.int/data/gho/data/themes/mortality-and-global-health-estimates/global-health-estimates-leading-causes-of-dalys), representing a huge economic health burden and causing untold suffering for patients and families (Wu et al., 2005). Given the societal and economic costs of psychotic illness and the limited efficacy of current treatment options, it is of high strategic importance that we improve our understanding of the neurobiological mechanisms that underlie schizophrenia spectrum disorders, to inform better detection and stratification and treatment development. Over recent decades a key line of research in schizophrenia and psychosis populations has been to investigate neuroanatomical markers relative to healthy control groups. Numerous Magnetic Resonance Imaging (MRI) studies investigating brain structure in schizophrenia and psychosis risk populations (i.e. clinical risk, familial risk, and schizotypy) have been funded in the UK and internationally, progressing our understanding of the neuroanatomical basis of schizophrenia and of these psychosis risk populations. The first neuroimaging anatomical investigation of patients with schizophrenia, using CT scans, was published in 1976 (Johnstone et al., 1976), and at the time of writing the current paper, a PubMed search using the terms ‘*schizophrenia MRI (volume or thickness)*’ show that 3,203 publications reporting the neuroanatomical basis of schizophrenia using MRI were published between 1986 and August 2024. Many structural MRI studies have also been published in first episode and psychosis risk populations. However, the majority of these MRI studies, particularly older studies, have small sample sizes in the range of 10 – 100 (Button et al., 2013) due to the high costs of MRI, alongside difficulties recruiting patients with schizophrenia and other psychotic illnesses at a single centre. Whilst the field has advanced significantly in recent decades, there have been some reproducibility issues in the literature (Marek et al., 2022) hampering our ability to develop a definitive model of the neuroanatomical basis of schizophrenia, and the illness’s developmental trajectory.

In an attempt to address this problem, there have recently been a number of notable large, international multisite MRI initiatives and consortia in focusing on individuals at high clinical risk for psychosis or those with a diagnosis of schizophrenia such as EU-GEI (Modinos et al., 2020), PRONIA (Rosen et al., 2021), PSYSCAN (Tognin et al., 2020), PHENOM (Chand et al., 2020). Additionally, there have been meta and mega analysis of existing data by the ENIGMA consortium (ENIGMA Clinical High Risk for Psychosis Working Group, 2021; Lamsma et al., 2024; van Erp et al., 2018). Whilst these have significantly advanced the field by improving our understanding of the anatomical changes seen in schizophrenia and psychosis risk populations, as well as addressing power issues, these consortia studies mostly have an over-representation of samples from European, Australian, and North American sites. The paucity of samples from diverse ethnoracial groups (Fonseca et al., 2021) prevents conclusion as to whether patients from different backgrounds may show distinct patterns of neural markers (Fearon et al., 2006; Hutchinson et al., 1999; Kalra et al., 2012; Lim et al., 2011; Suhail & Cochrane, 2002). Samples of at risk populations and schizotypy are underrepresented in existing multi-centre consortia. Importantly, there is also a major lack of open-access neuroimaging data in psychosis: those that exist (e.g. http://schizconnect.org/) are severely limited in terms of size, global reach, and what data and support is available to researchers. Although mechanisms exist within the ENIGMA consortia to share data beyond consortia members, some consortia initiatives often restrict data access to consortia members only, making it difficult for aspiring researchers outside these consortia to conduct well-powered analyses. Thus, despite the considerable cost and effort associated with the acquisition and analyses of MRI datasets in schizophrenia spectrum disorder, first episode psychosis, and psychosis risk populations, clarity around the role of key brain regions and how these evolve across the disease trajectory, and their link to symptoms, is still lacking (Alnæs et al., 2019; Honea et al., 2005). Whilst ENIGMA includes consortia focusing on clinical high risk (ENIGMA Clinical High Risk Working Group et al., 2021) and schizotypy (ENIGMA Schizotypy Working Group et al., 2022) populations, there has been a tendency for both meta- and mega-analyses to focus on one disease stage only (typically chronic, medicated patients), this has limited our ability to draw inferences regarding underlying stage-specific neuroanatomical changes and mechanisms and understand how these contribute to disease progression (Keshavan et al., 2020). Finally, all meta-analyses, and even the majority of mega-analyses, have relied on summary data from contributing studies/centres. Furthermore, whilst there have been major efforts to harmonise feature level data (e.g. cortical thickness and surface area measures) across participating centres, voxel level analyses have been more difficult to achieve until more recently (Si et al., 2024).

Another significant obstacle to progress in the field is the limited access to MRI datasets within schizophrenia and psychosis populations, which remains largely confined to a select group of researchers. In the UK, for instance, only a few centres have collected and published MRI data on schizophrenia populations. The study of psychosis risk cohorts is even more concentrated, with research primarily conducted at centres in London, Cambridge, Birmingham (for clinical high risk), and Edinburgh (for familial high risk). This situation underscores the need to promote greater equity in scientific research, aligning with the UK government’s “levelling up” agenda. The current inequity has hindered the field’s ability to pursue essential new research, address existing knowledge gaps, and resolve inconsistencies. Additionally, early career researchers often face challenges in accessing these MRI datasets, limiting their opportunities for development and research, which in turn weakens overall research capacity in this critical area.

First, although the prevalence of schizophrenia is uniform globally (Jongsma et al., 2018), there are subtle variations in the illness characteristics, including long-term outcomes, across geographies and ethnocultural spaces (see (Kalra et al., 2012) for review). Second, many brain-based investigations into the pathogenesis of schizophrenia have been conducted in the global North and other developed countries, while a majority of the schizophrenia populace resides in the global South (Kalra et al., 2012). Lastly, there is a growing literature on how ethnicity (encompassing genetic, linguistic, cultural, and environmental factors) can impact brain structure and function (Gong et al., 2015; Strawbridge et al., 2018).

## 2. Psychosis Shared MRI Data Resource (Psy-ShareD) & Methods

### 2.1 Agents

For clarity going forward here we define terms for the following agents and groups which are used in the sections below.

- *The Psy-ShareD Partnership* comprises *Data Contributors* and *Team Members*.
- *Data Contributors* are those agents that have contributed MRI and linked clinical, demographic and cognitive data to Psy-ShareD.
- *Team Members* are those agents that are co-investigators and project staff. *Team Members* can also be *Data Contributors*.
- *Data Users* are those agents that access Psy-ShareD data for analyses and publication. *Data Users* can also be *Data Contributors* and *Team Members*.

### 2.2 Aims and Objectives

The *Psy-ShareD Partnership* is funded by the UK Medical Research Council (MR/X010651/1) with the remit to combine high quality, pre-existing ‘legacy’ structural MRI datasets, with linked clinical and cognitive data, into one free-to-access resource. Psy-ShareD is suitable for a-priori hypothesis testing, exploration for novel hypothesis generation, and methodological training. Specifically, the *Psy-ShareD Partnership* brings together pre-existing structural MRI scans in people with schizophrenia (SCZ), first episode psychosis (FEP), clinical high risk (CHR) for psychosis, schizotypy (SZT), and healthy control (HC) populations. The resource also includes MRI data from people diagnosed with mood disorders, notably major depressive disorder (MDD) and bipolar disorder (BPD). The latter populations, although not belonging to the psychosis spectrum, are diagnostic groups we plan to expand in the future (see section 4). The resource will also contain measures of childhood trauma (CT) in both clinical and healthy control populations potentially allowing categorical and correlational type analyses examining effects of early experiences on neuroanatomy. Overall, Psy-ShareD will allow comparisons amongst various illness phenotypes, and across illness stages, enhancing our capacity to understand both the neuroanatomical basis *and their* trajectory of schizophrenia and psychosis. A full list of available datasets can be viewed on the Psy-ShareD website (https://psyshared.com/Data.html).

To summarise, the primary objectives of the *Psy-ShareD Partnership* are as follows:

1. Build a sustainable free-to-access structural MRI data repository from pre-existing MRI datasets in Schizophrenia, First Episode Psychosis, Clinical High Risk, Schizotypy, Bipolar, Major Depressive Disorder, and HC participants, and include within the database linked demographic, clinical, and IQ data. Neuropsychological, medication, treatment responsiveness, and functional outcome measures will also be included where available.
2. Seek out further MRI datasets, particularly those acquired in non-Western populations.
3. Organise and curate all Psy-ShareD datasets and catalogue these clearly, via Kings College London (KCL) FigShare (https://kcl.figshare.com) and the Psy-ShareD Website (https://psyshared.com).
4. Undertake and publish a series of validation and proof-of-concept analyses using Psy-ShareD datasets to demonstrate reliability and feasibility of use.

### 2.3 Data Transfer

Contributing centres will share raw T1 images after defacing and removing any personally identifying information. This allows *Psy-ShareD* to conduct rigorous harmonisation procedures based on the raw data while safeguarding participants’ privacy and better accounting for variability between sites, and providing users with a higher level of reliability and power.

All Psy-ShareD partner institutions that share data complete a Data Sharing Agreement (DSA). More information about the Psy-ShareD Data Sharing Agreement process are provided on our website (https://psyshared.com/Datacontribution.html). Once a DSA is established with the contributing partner/institution, and prior to T1 and linked data transfer, the *Psy-ShareD Team* provide the transferring sites with scripts and instructions for data de-identification (de-facing) and full anonymisation, in accordance with the Psy-ShareD Data Transfer Protocol (Figure 1). A script is also provided to produce unique Psy-ShareD ID codes for each MRI T1 scan transferred from the contributing site. These ID codes also link MRI and corresponding clinical, demographic, and cognitive data. De-identification and anonymisation scripts are available via the Psy-ShareD website. Once data preparation steps are completed, data is transferred via BitBox (https://www.bitbox-imaging.com). BitBox (Easmin et al., 2022) was developed for the purpose of imaging and clinical data transfer within multi-site studies. BitBox is used to transfer medical imaging data from external and independent sites into the KCL neuroimaging network, securely and in accordance with GDPR regulations.

**Figure 1:**
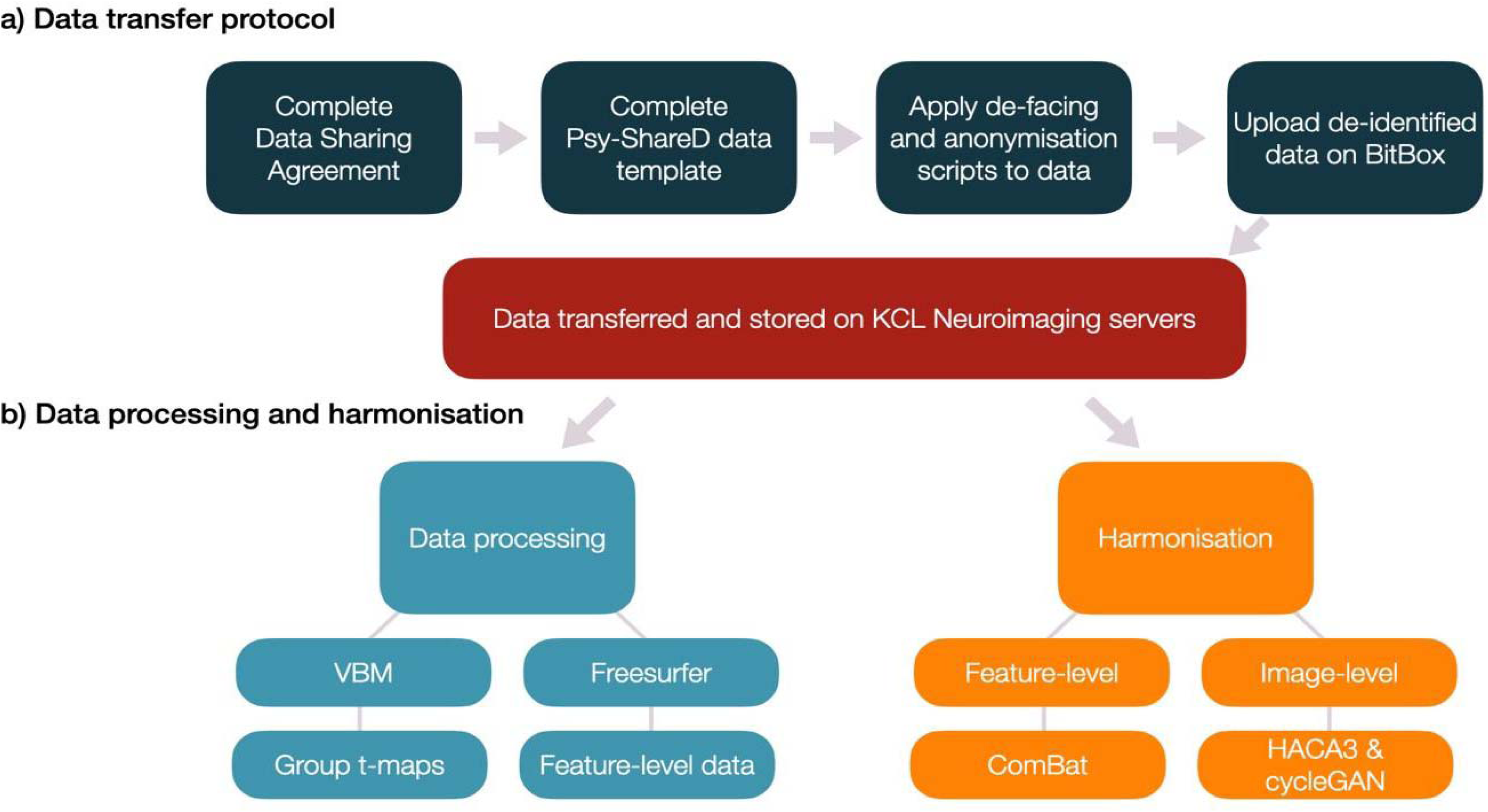
Schematic showing Psy-ShareD procedures for (a) data transfer and preparation steps taken by the Data Contributors (dark blue) prior to storage (red), (b) data processing (light blue) and harmonisation (orange) steps and results carried out by the Psy-ShareD team.

### 2.4 Data harmonisation

The T1 MRI data in Psy-ShareD were acquired at different sites using various T1 acquisition sequences (e.g. SPGR, ADNI-GO, MPRAGE). To account for different acquisition parameters used across sites, we harmonised the data using two types of methods: i) feature-level and ii) image-level harmonisation. To make available feature-level data including cortical and subcortical volumes, cortical thickness, and surface area data, we have used FreeSurfer version 7.3.2 (https://surfer.nmr.mgh.harvard.edu/). We then used the neuroHarmonize package (https://github.com/rpomponio/neuroHarmonize; Pomponio et al., 2019), to harmonise feature level MRI data by removing unwanted variation induced by scanner differences such as differences in acquisition parameters, field strength and manufacturer, while preserving biological variability between individuals using an empirical Bayes framework (Fortin et al., 2017, 2018; Johnson et al., 2007; Pomponio et al., 2019). Since this harmonisation is sample-dependent, Psy-ShareD tools are available to aid with feature level harmonisation. This allows users to harmonise feature-level MRI data, effectively removing unwanted variation due to scanner differences while preserving the biological variability between individuals, using an empirical Bayes framework (Fortin et al., 2017, 2018; Johnson et al., 2007; Pomponio et al., 2019). Site harmonisation with the neuroHarmonize package can be applied to all or a sub-set of the datasets in Psy-ShareD as per users’ requirements. Two advantages of neuroHarmonize over other methods (e.g. using site as a covariate in statistical models) are that it improves the removal of scanner effects in datasets with small sample sizes and does not make assumptions about the neuroimaging technique being used (Radua et al., 2020).

A unique feature of Psy-ShareD is the use of image-level harmonisation to T1 data, which produces whole brain images where site-specific information is removed, therefore allowing analyses such as voxel-based morphometry (VBM; Figure 2). We are employing two distinct methods to achieve this: HACA3 (Zuo et al., 2023) and 3D CycleGAN (Roca et al., 2023). HACA3 is a deep-learning method with an “encoder-attention-decoder” architecture (Zuo et al., 2023). In its simpler form, it takes two types of input images: a target image, and an original input image to be harmonised towards the target image. It then encodes the anatomy, contrast, and image artifact from the inputs. The method creates a synthetic image with the anatomical information from the original input image, but with the target image’s contrast, while removing artifacts (Zuo et al., 2023). By harmonising an image to a target (Figure 2A), original contrast values from the original images are moved to values in the range of a target image (Figure 2B), whilst maintaining its anatomical features (Figure 2C). Full data validation work using these harmonisation processes is underway and will be reported separately.

**Figure 2:**
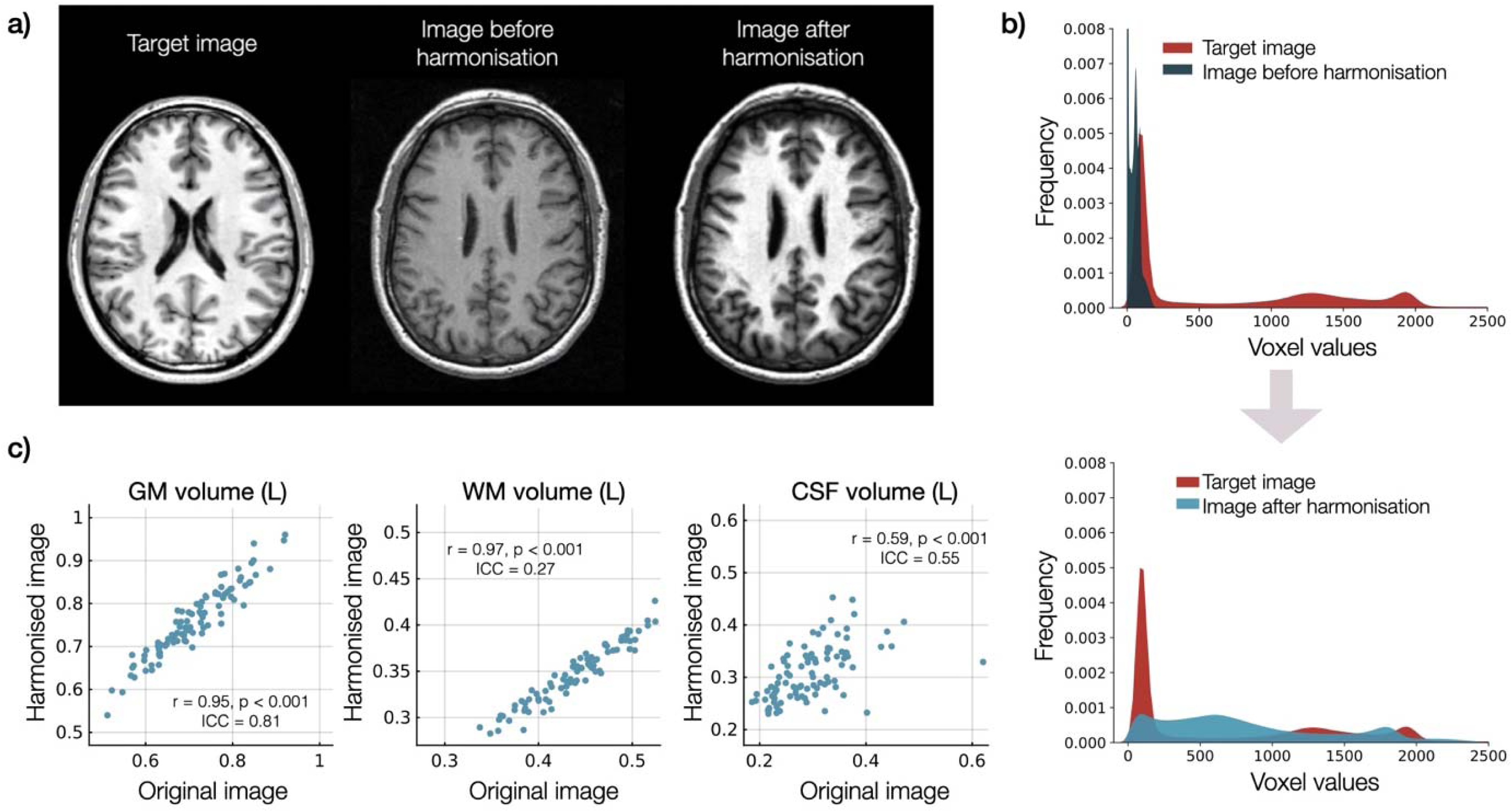
Example of harmonisation. a) Single subject image that was harmonised towards a target image. b)Voxel value frequency plot showing the values of voxels of the example image shown on (a) before and after it was harmonised towards a target image, c) correlation plots showing the volumes of white matter (left), grey matter (centre) and cerebrospinal fluid (right) after segmenting several original images and their harmonised version (HACA3 image). Each plot also depicts the correlation coefficient and corresponding p-value, as well as the intra-class correlation coefficient (ICC(A,1) or two-way random absolute agreement measure; McGraw & Wong 1996) between the values obtained with both images.

The *Psy-ShareD Team* is also implementing CycleGAN (Cycle Generative Adversarial Network) architecture (Zhu et al., 2017), which features two generator-discriminator pairs (labelled as G1, D1, G2, D2). The generator produces synthetic images that closely resemble the real dataset distribution, while the discriminator distinguishes between synthetic and real images. CycleGAN incorporates a cycle-consistency constraint in its loss function to ensure that unpaired image translation and content (anatomical structure) preservation are achieved. This model is particularly beneficial for image level harmonisation as it requires no supervision during the training process. Since no ground truth is required for training, the output of the second GAN (Generative Adversarial Network) must correspond only with the input of the first GAN. This means that there is no need for acquiring scans from the same participant at each site (Cackowski et al., 2021). Given that the original study focused on a 2D deep learning framework, we used the 3D framework of CycleGAN proposed by Roca et al., 2023. This adaptation uses U-Net generators and PatchGAN discriminators with 3D convolutions to process entire 3D MRI scans.

### 2.5 Data description

All MRI T1 data are linked to anonymised demographic and clinical data as detailed in our data catalogues available via KCL FigShare (https://kcl.figshare.com). All datasets include information about participants’ age and sex. Ethnoracial data, educational level, and handedness are also available in many datasets. All datasets include caseness and several datasets also include data for illness duration/onset, and medication status. Symptom severity in SCZ and FEP populations were derived from the Positive and Negative Symptom Scale (PANSS; (Kay et al., 1987)) and the Scale for the Assessments of Positive and Negative Symptoms (SAPS/SANS; (Andreasen, 1984)). Cohorts of individuals at high risk for psychosis were assessed for caseness and severity using the Comprehensive Assessment for an At-Risk Mental States (CAARMS; Yung et al., 2005) and with SIPS/SOP (McGlashan et al., 2001) for some datasets. Global functioning (General Assessment of Functioning; GAF (Hall, 1995)) and measures for depression and anxiety symptoms are also available in several datasets. In non-clinical risk groups (SZT and CT) a range of sub-clinical measures are provided where available, such as Oxford-Liverpool Inventory for Feelings and Experiences (Mason & Claridge, 2006), Schizotypal Personality Questionnaire (Raine, 1991) and Childhood Trauma Questionnaire (Bernstein et al., 2003). Cognitive and IQ data are also available in several datasets assessing intellectual function, working memory and executive function, and verbal learning (see data catalogues).

## 3. Psy-ShareD Data Management: Access and Storage

### 3.1 Data Access and Storage

Data catalogues are available through KCL FigShare (https://kcl.figshare.com) and the Psy-ShareD website (https://psyshared.com/Data.html). Data catalogues contain information about each dataset, including, caseness, sample size, data formats, demographics, clinical, medication, and cognitive variables.

MRI T1 and linked clinical/demographic/cognitive data will be stored separately on the KCL Neuroimaging Network and will not be directly accessible to users. Members of the Psy-ShareD Partnership act as custodians of the data contained within the resource. Potential Data Users can access data contained in the Psy-ShareD database by submitting a data access request using a short Data Access Form available on the Psy-ShareD website. The data access procedure is shown in Figure 3. The data access procedure is required to track and monitor access and allow data contributors whose datasets are requested to provide approval.

**Figure 3:**
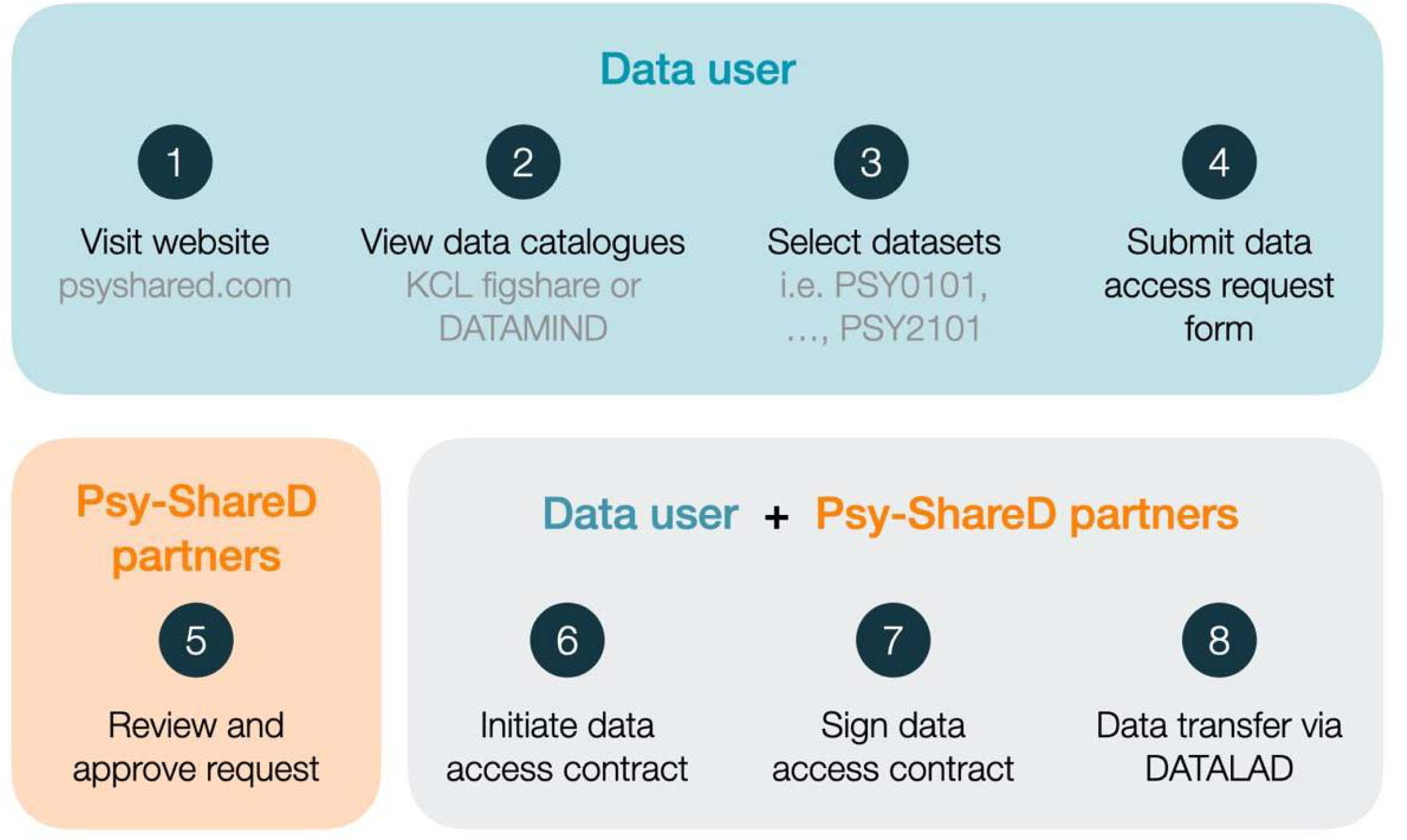
Psy-ShareD Data Access Procedure

Once a project is approved, after data access request, data access will be managed via DataLad and GIN (G-Node Infrastructure). DataLad (https://www.datalad.org/) is an open-source data management tool that tracks data and ensures reproducibility. On the other hand, GIN (https://gin.g-node.org/) is a free and open platform for data storage and distribution, allowing data to be accessed and managed from various locations while keeping it synchronised and backed up. After the data is prepared with DataLad, it will be shared through the GIN application. This combination supports dataset versioning and provides controlled access for collaborators.

All Psy-ShareD Data Users will be required to adhere to the Memorandum of Understanding (MoU) which can be accessed via the Psy-ShareD website. The Psy-ShareD MoU provides details and instructions for Psy-ShareD *Data Users* about publication, authorship, and open access data arrangements.

### 3.2 Data Use and Publication Policy

Full details are available in the Psy-ShareD Memorandum of Understanding (MoU) document (see website). Briefly, we stipulate that all outputs that use Psy-ShareD datasets include Data Contributors as co-authors. In addition, the *Psy-ShareD Partnership* will be listed on the author line of all publications using data from the Psy-ShareD resource. As such, all publications and citations resulting from Psy-ShareD will be linked to the respective Data Contributors. Specific publication policies (i.e. conference abstracts, posters, symposia) are listed in the Psy-ShareD MoU document.

### 3.3 Tools, Training and Open Access

For partners that contribute data, instructions and tools for data preparation and transfer are available free upon request from the Psy-ShareD team. Tools for T1 image level harmonisation using HACA3 and CycleGan and using ComBat for feature level harmonisation will also be available via the Psy-ShareD website. To facilitate usage of Psy-ShareD data, the Psy-ShareD Partnership will develop and provide supporting material and workshops for potential users. These will include open-access training resources for data organisation and analysis. Data sharing plans are fully in line with the UK Medical Research Council’s data sharing policy and Open Science Framework principles (https://osf.io) that promote and support networking and partnership activities, enabling knowledge sharing and open access to data across institutions.

## 4. Future Directions and Psy-ShareD Phase II

Psy-ShareD is a growing resource and new datasets from around the world are continually sought and being added on an ongoing basis. Currently, MRI datasets are available from sites in the UK, Europe, South & Central America, India, and Japan. Further, data sharing agreements are currently in progress with sites in North America and Australia as the resource continues to expand. We also plan to expand Psy-ShareD trans diagnostically. The resource already includes MRI scans from patients diagnosed with bipolar disorder and MDD and it is planned that MRI data from cohorts with other DSM-5 disorders will be added to the resource in the future. It is also anticipated that, going forward, data from other MRI modalities can be added and linked to Psy-ShareD T1 data, i.e., resting state functional MRI, 1H-Magnetic Resonance Spectroscopy. In summary, future directions and objectives are as follows:

▯ General expansion to new sites, especially focusing on enhancing geographical and ethnoracial diversity.
▯ Expanding beyond T1 datasets to multimodal MRI (i.e. resting state fMRI, MRS).
▯ Increasing the range of diagnoses of future clinical samples.
▯ Inclusion of genetic & omics data (inflammatory markers etc.) where possible.
▯ Development of clinical tools, using Psy-ShareD data.

We are exploring methodological approaches that will allow Psy-ShareD data to be used for clinical applications, e.g., clustering and neurobiological stratification (Lalousis et al., 2022) and normative modelling of brain morphometry (Haas et al. 2024) approaches. Finally, we will link our data catalogues to DATAMIND (https://datamind.org.uk) and we are exploring the possibility of also linking to HDRUK (https://www.hdruk.ac.uk).

## Data Availability

All data produced are available online at
https://psyshared.com/Home.html

https://psyshared.com/Home.html

## References

Alnæs, D., Kaufmann, T., van der Meer, D., Córdova-Palomera, A., Rokicki, J., Moberget, T., Bettella, F., Agartz, I., Barch, D. M., Bertolino, A., Brandt, C. L., Cervenka, S., Djurovic, S., Doan, N. T., Eisenacher, S., Fatouros-Bergman, H., Flyckt, L., Di Giorgio, A., Haatveit, B., … Westlye, L. T. (2019). Brain Heterogeneity in Schizophrenia and Its Association With Polygenic Risk. JAMA Psychiatry, 76(7), 739–748. 10.1001/jamapsychiatry.2019.0257

Andreasen, N. C. (1984). Scale for the Assessment of Positive Symptoms. Psychiatrie & Psychobiologie. 10.1037/t48377-000

Bernstein, D. P., Stein, J. A., Newcomb, M. D., Walker, E., Pogge, D., Ahluvalia, T., Stokes, J., Handelsman, L., Medrano, M., Desmond, D., & Zule, W. (2003). Development and validation of a brief screening version of the Childhood Trauma Questionnaire. Child Abuse & Neglect, 27(2), 169–190. 10.1016/s0145-2134(02)00541-0

Button, K.S., Ioannidis JP, Mokrysz C, Nosek BA, Flint J, Robinson ES, Munafò MR. Power failure: why small sample size undermines the reliability of neuroscience. Nat Rev Neurosci. 2013 May;14(5):365–76. doi: 10.1038/nrn3475. Epub 2013 Apr 10. Erratum in: Nat Rev Neurosci. 2013 Jun;14(6):451. PMID: 23571845.

Cackowski, S., Barbier, E. L., Dojat, M., & Christen, T. (2021). comBat versus cycleGAN for multi-center MR images harmonization. https://openreview.net/forum?id=cbJD-wMlJK0

Chand, G. B., Dwyer, D. B., Erus, G., Sotiras, A., Varol, E., Srinivasan, D., Doshi, J., Pomponio, R., Pigoni, A., Dazzan, P., Kahn, R. S., Schnack, H. G., Zanetti, M. V., Meisenzahl, E., Busatto, G. F., Crespo-Facorro, B., Pantelis, C., Wood, S. J., Zhuo, C., … Davatzikos, C. (2020). Two distinct neuroanatomical subtypes of schizophrenia revealed using machine learning. Brain: A Journal of Neurology, 143(3), 1027–1038. 10.1093/brain/awaa025

Easmin, R., Nordio, G., Giacomel, A., Turkheimer, F., Williams, S., & Veronese, M. (2022). Bitbox: A Cloud-based data sharing solution for medical images. Annual International Conference of the IEEE Engineering in Medicine and Biology Society. IEEE Engineering in Medicine and Biology Society. Annual International Conference, 2022, 2712–2715. 10.1109/EMBC48229.2022.9871689

ENIGMA Clinical High Risk for Psychosis Working Group. (2021). Association of Structural Magnetic Resonance Imaging Measures With Psychosis Onset in Individuals at Clinical High Risk for Developing Psychosis: An ENIGMA Working Group Mega-analysis. JAMA Psychiatry, 78(7), 753–766. 10.1001/jamapsychiatry.2021.0638.

ENIGMA Clinical High Risk for Psychosis Working Group (2022). Association of Structural Magnetic Resonance Imaging Measures With Psychosis Onset in Individuals at Clinical High Risk for eveloping Psychosis: An ENIGMA Working Group Mega-analysis. JAMA Psychiatry. 2021 Jul 1;78(7):753–766. doi: 10.1001/jamapsychiatry.2021.0638.

Fearon, P., Kirkbride, J. B., Morgan, C., Dazzan, P., Morgan, K., Lloyd, T., Hutchinson, G., Tarrant, J., Fung, W. L. A., Holloway, J., Mallett, R., Harrison, G., Leff, J., Jones, P. B., & Murray, R. M. (2006). Incidence of schizophrenia and other psychoses in ethnic minority groups: Results from the MRC AESOP Study. Psychological Medicine, 36(11), 1541–1550. 10.1017/S0033291706008774

Fonseca L, Sena BF, Crossley N, Lopez-Jaramillo C, Koenen K, Freimer NB, Bressan RA, Belangero SI, Santoro ML, Gadelha A. Diversity matters: opportunities in the study of the genetics of psychotic disorders in low- and middle-income countries in Latin America. Braz J Psychiatry. 2021 Nov-Dec;43(6):631–637. doi: 10.1590/1516-4446-2020-1240. PMID: 33237255; PMCID:

Fortin, J.-P., Cullen, N., Sheline, Y. I., Taylor, W. D., Aselcioglu, I., Cook, P. A., Adams, P., Cooper, C., Fava, M., McGrath, P. J., McInnis, M., Phillips, M. L., Trivedi, M. H., Weissman, M. M., & Shinohara, R. T. (2018). Harmonization of cortical thickness measurements across scanners and sites. NeuroImage, 167, 104–120. 10.1016/j.neuroimage.2017.11.024

Fortin, J.-P., Parker, D., Tunç, B., Watanabe, T., Elliott, M. A., Ruparel, K., Roalf, D. R., Satterthwaite, T. D., Gur, R. C., Gur, R. E., Schultz, R. T., Verma, R., & Shinohara, R. T. (2017). Harmonization of multi-site diffusion tensor imaging data. NeuroImage, 161, 149–170. 10.1016/j.neuroimage.2017.08.047

Ge, R., Yu, Y., Qi, Y. X., Fan, Y., Chen, S., Gao, C., Haas, S. S., New, F., Boomsma, D. I., Brodaty, H., Brouwer, R. M., Buckner, R., Caseras, X., Crivello, F., Crone, E. A., Erk, S., Fisher, S. E., Franke, B., Glahn, D. C., … Yu, K. (2024). Normative modelling of brain morphometry across the lifespan with CentileBrain: Algorithm benchmarking and model optimisation. The Lancet Digital Health, 6(3), e211–e221. 10.1016/S2589-7500(23)00250-9

Gong, Q., Dazzan, P., Scarpazza, C., Kasai, K., Hu, X., Marques, T. R., Iwashiro, N., Huang, X., Murray, R. M., Koike, S., David, A. S., Yamasue, H., Lui, S., & Mechelli, A. (2015). A Neuroanatomical Signature for Schizophrenia Across Different Ethnic Groups. Schizophrenia Bulletin, 41(6), 1266–1275. 10.1093/schbul/sbv109

Haas, S. S., Ge, R., Agartz, I., Amminger, G. P., Andreassen, O. A., Bachman, P., Baeza, I., Choi, S., Colibazzi, T., Cropley, V. L., de la Fuente-Sandoval, C., Ebdrup, B. H., Fortea, A., Fusar-Poli, P., Glenthøj, B. Y., Glenthøj, L. B., Haut, K. M., Hayes, R. A., Heekeren, K., … Frangou, S. (2023). Normative modeling of brain morphometry in Clinical High-Risk for Psychosis. bioRxiv, 2023.01.17.523348. 10.1101/2023.01.17.523348

Hall, R. C. (1995). Global assessment of functioning. A modified scale. Psychosomatics, 36(3), 267–275. 10.1016/S0033-3182(95)71666-8

Honea, R., Crow, T. J., Passingham, D., & Mackay, C. E. (2005). Regional Deficits in Brain Volume in Schizophrenia: A Meta-Analysis of Voxel-Based Morphometry Studies. American Journal of Psychiatry, 162(12), 2233–2245. 10.1176/appi.ajp.162.12.2233

Hutchinson, G., Takei, N., Sham, P., Harvey, I., & Murray, R. M. (1999). Factor analysis of symptoms in schizophrenia: Differences between White and Caribbean patients in Camberwell. Psychological Medicine, 29(3), 607–612. 10.1017/S0033291799008430

Johnson, W. E., Li, C., & Rabinovic, A. (2007). Adjusting batch effects in microarray expression data using empirical Bayes methods. Biostatistics, 8(1), 118–127. 10.1093/biostatistics/kxj037

Johnstone, E., Frith, C. D., Crow, T. J., Husband, J., & Kreel, L. (1976). Cerebral ventricular size and cognitive impairment in chronic schizophrenia. The Lancet, 308(7992), 924–926. 10.1016/S0140-6736(76)90890-4

Jongsma, H. E., Gayer-Anderson, C., Lasalvia, A., Quattrone, D., Mulè, A., Szöke, A., Selten, J.-P., Turner, C., Arango, C., Tarricone, I., Berardi, D., Tortelli, A., Llorca, P.-M., de Haan, L., Bobes, J., Bernardo, M., Sanjuán, J., Santos, J. L., Arrojo, M., … European Network of National Schizophrenia Networks Studying Gene-Environment Interactions Work Package 2 (EU-GEI WP2) Group. (2018). Treated Incidence of Psychotic Disorders in the Multinational EU-GEI Study. JAMA Psychiatry, 75(1), 36–46. 10.1001/jamapsychiatry.2017.3554

Kalra, G., Bhugra, D., & Shah, N. (2012). Cultural aspects of schizophrenia. International Review of Psychiatry, 24(5), 441–449. 10.3109/09540261.2012.708649

Kay, S. R., Fiszbein, A., & Opler, L. A. (1987). The Positive and Negative Syndrome Scale (PANSS) for Schizophrenia. Schizophrenia Bulletin, 13(2), 261–276. 10.1093/schbul/13.2.261

Keshavan, M. S., Collin, G., Guimond, S., Kelly, S., Prasad, K. M., & Lizano, P. (2020). Neuroimaging in schizophrenia. Neuroimaging Clinics of North America, 30(1), 73–83. 10.1016/j.nic.2019.09.007

Lalousis, P. A., Schmaal, L., Wood, S. J., Reniers, R. L. E. P., Barnes, N. M., Chisholm, K., Griffiths, S. L., Stainton, A., Wen, J., Hwang, G., Davatzikos, C., Wenzel, J., Kambeitz-Ilankovic, L., Andreou, C., Bonivento, C., Dannlowski, U., Ferro, A., Lichtenstein, T., Riecher-Rössler, A., … Upthegrove, R. (2022). Neurobiologically Based Stratification of Recent-Onset Depression and Psychosis: Identification of Two Distinct Transdiagnostic Phenotypes. Biological Psychiatry, 92(7), 552–562. 10.1016/j.biopsych.2022.03.021

Lamsma, J., Raine, A., Kia, S. M., Cahn, W., Arold, D., Banaj, N., Barone, A., Brosch, K., Brouwer, R., Brunetti, A., Calhoun, V. D., Chew, Q. H., Choi, S., Chung, Y.-C., Ciccarelli, M., Cobia, D., Cocozza, S., Dannlowski, U., Dazzan, P., … Nickl-Jockschat, T. (2024). Structural brain abnormalities and aggressive behaviour in schizophrenia: Mega-analysis of data from 2095 patients and 2861 healthy controls via the ENIGMA consortium. medRxiv: The Preprint Server for Health Sciences, 2024.02.04.24302268. 10.1101/2024.02.04.24302268

Lim, C. S., Mythily Subramaniam, Lye Yin Poon, Siow Ann Chong, & Swapna Verma. (2011). Cross-ethnic differences in severity of symptomatology of individuals with first-episode schizophrenia spectrum disorder. Early Intervention in Psychiatry, 242–248.

Lopez, A. D., Mathers, C. D., Ezzati, M., Jamison, D. T., & Murray, C. J. (Eds.). (2006). Global Burden of Disease and Risk Factors. The International Bank for Reconstruction and Development / The World Bank. http://www.ncbi.nlm.nih.gov/books/NBK11812/

Marek, S., Tervo-Clemmens, B., Calabro, F. J., Montez, D. F., Kay, B. P., Hatoum, A. S., Donohue, M. R., Foran, W., Miller, R. L., Hendrickson, T. J., Malone, S. M., Kandala, S., Feczko, E., Miranda-Dominguez, O., Graham, A. M., Earl, E. A., Perrone, A. J., Cordova, M., Doyle, O., … Dosenbach, N. U. F. (2022). Reproducible brain-wide association studies require thousands of individuals. Nature, 603(7902), Article 7902. 10.1038/s41586-022-04492-9

Mason, O., & Claridge, G. (2006). The Oxford-Liverpool Inventory of Feelings and Experiences (O-LIFE): Further description and extended norms. Schizophrenia Research, 82(2), 203–211. 10.1016/j.schres.2005.12.845

McGlashan TH, Miller TJ, Woods SW, Hoffman RE, Davidson L. A scale for the assessment of prodromal symptoms and states. In: Miller TJ, Mednick SA, McGlashan TT, Liberger J, Johannessen JO, editors. Early Intervention in Psychotic Disorders. Dordrecht, The Netherlands: Kluwer Academic Publishers; 2001. pp. 135–149

McGraw, K. O., & Wong, S. P. (1996). Forming inferences about some intraclass correlation coefficients. Psychological methods, 1(1), 30.

Modinos, G., Kempton, M. J., Tognin, S., Calem, M., Porffy, L., Antoniades, M., Mason, A., Azis, M., Allen, P., Nelson, B., McGorry, P., Pantelis, C., Riecher-Rössler, A., Borgwardt, S., Bressan, R., Barrantes-Vidal, N., Krebs, M.-O., Nordentoft, M., Glenthøj, B., … EU-GEI High Risk Study Group. (2020). Association of Adverse Outcomes With Emotion Processing and Its Neural Substrate in Individuals at Clinical High Risk for Psychosis. JAMA Psychiatry, 77(2), 190–200. 10.1001/jamapsychiatry.2019.3501

Radua, J., Vieta, E., Shinohara, R., Kochunov, P., Quidé, Y., Green, M. J., Weickert, C. S., Weickert, T., Bruggemann, J., Kircher, T., Nenadić, I., Cairns, M. J., Seal, M., Schall, U., Henskens, F., Fullerton, J. M., Mowry, B., Pantelis, C., Lenroot, R., … van Erp, T. (2020). Increased power by harmonizing structural MRI site differences with the ComBat batch adjustment method in ENIGMA. NeuroImage, 218, 116956. 10.1016/j.neuroimage.2020.116956

Raine, A. (1991). The SPQ: A scale for the assessment of schizotypal personality based on DSM-III-R criteria. Schizophrenia Bulletin, 17(4), 555–564. 10.1093/schbul/17.4.555

Roca V, Kuchcinski G, Pruvo JP, Manouvriez D, Leclerc X, Lopes R. A three-dimensional deep learning model for inter-site harmonization of structural MR images of the brain: Extensive validation with a multicenter dataset. Heliyon. 2023 Dec 1;9(12).

Rosen, M., Betz, L. T., Schultze-Lutter, F., Chisholm, K., Haidl, T. K., Kambeitz-Ilankovic, L., Bertolino, A., Borgwardt, S., Brambilla, P., Lencer, R., Meisenzahl, E., Ruhrmann, S., Salokangas, R. K. R., Upthegrove, R., Wood, S. J., Koutsouleris, N., & Kambeitz, J. (2021). Towards Clinical Application of Prediction Models for Transition to Psychosis: A Systematic Review and External Validation Study in the PRONIA Sample. Neuroscience and Biobehavioral Reviews, 125, 478–492. 10.1016/j.neubiorev.2021.02.032

Si, S., Bi, A., Yu, Z., See, C., Kelly, S., Ambrogi, S., Arango, C., Baeza, I., Banaj, N., Berk, M., Castro-Fornieles, J., Crespo-Facorro, B., Crouse, J. J., Díaz-Caneja, C. M., Fett, A.-K., Fortea, A., Frangou, S., Goldstein, B. I., Hickie, I. B., … Kempton, M. J. (2024). Mapping gray and white matter volume abnormalities in early-onset psychosis: An ENIGMA multicenter voxel-based morphometry study. Molecular Psychiatry, 29(2), 496–504.

Strawbridge RJ, Ward J, Lyall LM, Tunbridge EM, Cullen B, Graham N, Ferguson A, Johnston KJA, Lyall DM, Mackay D, Cavanagh J, Howard DM, Adams MJ, Deary I, Escott-Price V, O’Donovan M, McIntosh AM, Bailey MES, Pell JP, Harrison PJ, Smith DJ. Genetics of self-reported risk-taking behaviour, trans-ethnic consistency and relevance to brain gene expression. Transl Psychiatry. 2018 Sep ;8(1):178. doi: 10.1038/s41398-018-0236-1. PMID: 30181555; PMCID: PMC6123450.

Suhail, K., & Cochrane, R. (2002). Effect of culture and environment on the phenomenology of delusions and hallucinations. The International Journal of Social Psychiatry, 48(2), 126–138. 10.1177/002076402128783181

Tognin, S., van Hell, H. H., Merritt, K., Winter-van Rossum, I., Bossong, M. G., Kempton, M. J., Modinos, G., Fusar-Poli, P., Mechelli, A., Dazzan, P., Maat, A., de Haan, L., Crespo-Facorro, B., Glenthøj, B., Lawrie, S. M., McDonald, C., Gruber, O., van Amelsvoort, T., Arango, C., … PSYSCAN Consortium. (2020). Towards Precision Medicine in Psychosis: Benefits and Challenges of Multimodal Multicenter Studies-PSYSCAN: Translating Neuroimaging Findings From Research into Clinical Practice. Schizophrenia Bulletin, 46(2), 432–441. 10.1093/schbul/sbz067

van Erp, T. G. M., Walton, E., Hibar, D. P., Schmaal, L., Jiang, W., Glahn, D. C., Pearlson, G. D., Yao, N., Fukunaga, M., Hashimoto, R., Okada, N., Yamamori, H., Bustillo, J. R., Clark, V. P., Agartz, I., Mueller, B. A., Cahn, W., de Zwarte, S. M. C., Hulshoff Pol, H. E., … Turner, J. A. (2018). Cortical Brain Abnormalities in 4474 Individuals With Schizophrenia and 5098 Control Subjects via the Enhancing Neuro Imaging Genetics Through Meta Analysis (ENIGMA) Consortium. Biological Psychiatry, 84(9), 644–654. 10.1016/j.biopsych.2018.04.023

Wu, E. Q., Birnbaum, H. G., Shi, L., Ball, D. E., Kessler, R. C., Moulis, M., & Aggarwal, J. (2005). The economic burden of schizophrenia in the United States in 2002. The Journal of Clinical Psychiatry, 66(9), 1122–1129. 10.4088/jcp.v66n0906

Yung, A. R., Yung, A. R., Pan Yuen, H., Mcgorry, P. D., Phillips, L. J., Kelly, D., Dell’olio, M., Francey, S. M., Cosgrave, E. M., Killackey, E., Stanford, C., Godfrey, K., & Buckby, J. (2005). Mapping the Onset of Psychosis: The Comprehensive Assessment of At-Risk Mental States. Australian & New Zealand Journal of Psychiatry, 39(11–12), 964–971. 10.1080/j.1440-1614.2005.01714.x

Zhu, J.-Y., Park, T., Isola, P., & Efros, A. A. (2017). Unpaired Image-to-Image Translation Using Cycle-Consistent Adversarial Networks. 2017 IEEE International Conference on Computer Vision (ICCV), 2242–2251. 10.1109/ICCV.2017.244

Zuo, L., Liu, Y., Xue, Y., Dewey, B. E., Remedios, S. W., Hays, S. P., Bilgel, M., Mowry, E. M., Newsome, S. D., Calabresi, P. A., Resnick, S. M., Prince, J. L., & Carass, A. (2023). HACA3: A unified approach for multi-site MR image harmonization. Computerized Medical Imaging and Graphics, 109, 102285. 10.1016/j.compmedimag.2023.102285

Zurita, M., Easmin, R., Bucci, S., Kumari, V., Evans, S., Kempton, M. J., & Allen, P. (2025) Evaluating harmonisation methods for Psy-ShareD T1w data: A comparative analysis. In preparation.

